# Trends in CD4 and viral load testing 2005 to 2018: Multi-cohort study of people living with HIV in Southern Africa

**DOI:** 10.1101/2020.03.09.20033423

**Authors:** Elizabeth Zaniewski, Cam Ha Dao Ostinelli, Frédérique Chammartin, Nicola Maxwell, Mary-Ann Davies, Jonathan Euvrard, Janneke van Dijk, Samuel Bosomprah, Sam Phiri, Frank Tanser, Nosisa Sipambo, Josephine Muhairwe, Geoffrey Fatti, Hans Prozesky, Robin Wood, Nathan Ford, Matthew P Fox, Matthias Egger

## Abstract

**Introduction:** WHO recommends a CD4 cell count before starting antiretroviral therapy (ART) to detect advanced HIV disease, and routine viral load (VL) testing following ART initiation to detect treatment failure. Donor support for CD4 testing has declined to prioritize access to VL monitoring. We examined trends in CD4 and VL testing among adults (≥15 years of age) starting ART in Southern Africa.

**Methods:** We analysed data from 14 HIV treatment programs with over 300 clinics in Lesotho, Malawi, Mozambique, South Africa, Zambia and Zimbabwe in years 2005-2018. We examined the frequency of CD4 and VL testing, the percentage of adults with CD4 or VL tests, and among those having a test, the percentage starting ART with advanced HIV disease (CD4 count <200 cells/mm^3^) or failing to suppress viral replication (>1000 HIV-RNA copies/ml) after ART initiation. We used mixed effect logistic regression to assess time trends adjusted for age and sex.

**Results:** Among 502,456 adults, the percentage with CD4 testing at ART initiation decreased from a high of 78.1% in 2008 to a low of 38.0% in 2017. The percentage starting with advanced HIV disease declined from 83.3% in 2005 to 23.5% in 2018. VL testing after starting ART varied; 61.0% of adults in South Africa and 10.7% in Malawi were tested but fewer than 2% were tested in the other four countries. The probability of having a CD4 cell count at ART start declined by 14% each year (odds ratio [OR] 0.86; 95% CI 0.86-0.86); the probability of advanced HIV disease declined by 20% per year (OR 0.80; 95% CI 0.80-0.81). The increase in VL testing after ART start was only modest (OR 1.06; 95% CI 1.05-1.06) and there was no evidence of a decrease in unsuppressed VL over time (OR 1.00; 95% CI 0.99-1.01).

**Conclusions:** CD4 cell counting declined over time, including testing at the start of ART, despite the fact that many patients still initiated ART with advanced HIV disease. Without CD4 testing and expanded VL testing many patients with advanced HIV disease and treatment failure may go undetected, threatening the effectiveness of ART in sub-Saharan Africa.

## Introduction

The World Health Organization (WHO) has recommended immediate initiation of antiretroviral therapy (ART) for all people living with HIV since 2015, regardless of CD4 cell count or clinical stage [1], but WHO still recommends CD4 cell counts at enrolment into HIV. The CD4 cell count is the best way to measure a patient’s immune status prior to starting ART or return to care following treatment interruption, and is the best predictor of disease progression and risk of death, especially among those with advanced HIV disease [2– 4]. CD4 cell count is used to guide a number of prophylactic and diagnostic interventions, including prioritizing testing for opportunistic infections, such as cryptococcosis or tuberculosis [5,6].

The primary benefit of ART is the suppression of HIV-1 viral replication [1,7,8]. Suppression of viral load (defined by WHO as ≤1000 HIV-RNA copies/ml) reduces morbidity and mortality among patients living with HIV and onward transmission of the virus [9]. It is the third of the UNAIDS global 90-90-90 and 95-95-95 targets launched in 2014, which aim to have 73% and 86%, of all people living with HIV virally suppressed by 2020 and 2030, respectively [10,11]. Since 2013, WHO has recommended viral load testing, rather than CD4 testing, as the preferred monitoring approach to detect treatment failure following ART initiation [12]. WHO guidelines were further refined in 2016 and currently recommend routine viral load testing six months after starting ART and yearly thereafter among patients stable on ART [1,7]. Patients with two consecutive unsuppressed viral load measurements (>1000 copies/ml) despite enhanced adherence counselling, taken within a three-month interval and at least six months after starting ART are considered to be in treatment failure [1,7,12]. Treatment failure indicates either some degree of drug resistance, or that ART has not been taken properly [13].

The President’s Emergency Plan for AIDS Relief (PEPFAR) [14] has been one of the largest financial commitments to combat HIV/AIDS, supporting more than 50 countries globally, including more than 20 countries in Africa [14,15]. In 2018, PEPFAR shifted testing priorities reducing the overall level of support for CD4 testing to prioritize access to viral load monitoring [16,17]. In this study, we examined longitudinal data from six countries in the International epidemiology Databases to Evaluate AIDS (IeDEA) Southern Africa region, all supported by PEPFAR since 2008, to assess trends in CD4 and viral load testing between 2005 and 2018.

## Methods

### Data sources

The IeDEA collaboration (www.iedea.org) is an international research and data consortium of HIV cohorts which has been funded by the National Institutes of Health since 2006 [18– 20]. IeDEA pools clinical and epidemiological data on people living with HIV under care in routine settings as a cost-effective way of generating large data sets to address research questions in HIV/AIDS treatment and care that cannot be answered by single cohorts [21]. The IeDEA Southern Africa region currently includes patient-level data on more than one million adults and children living with HIV from 17 large treatment programs across Lesotho, Malawi, Mozambique, South Africa, Zambia and Zimbabwe [22].

### Eligibility criteria

We included all CD4 cell count and viral load test measurements from adults aged ≥15 years who were ART-naÏve at enrolment and started ART between 2005 and 2018 at one of the HIV treatment programs currently supported by IeDEA Southern Africa. We only included public-access programs that collect adult CD4 and viral load test data using government (i.e. not privately funded) laboratory services. To ensure adequate time to capture test measurements we excluded adults who started ART less than one week before the date of database closure and adults who had less than six months of possible test measurement data available prior to ART start (e.g.. patients who started ART in the first six months of 2005).

### Outcomes

We assessed the number of CD4 cell count and viral load tests performed by exact calendar date from January 1, 2005 to December 31, 2018, irrespective of whether testing occurred before or after ART start, and calculated the ratio of CD4 cell count to viral load testing per year over time and stratified by country. We examined the percentage of adults with a CD4 cell count at ART start, the percentage of adults with any CD4 cell count up to six months before ART start, and, among those with a CD4 cell count, the percentage of adults starting ART with advance HIV disease by country and calendar year. We defined ART as a regimen of at least three antiretroviral drugs from two drug classes, and advanced HIV disease as a CD4 cell count <200 cells/mm^3^.

We defined CD4 cell count at ART start as a CD4 cell count taken within a time window of three months before and up to one week after ART start. In earlier years, it may have taken several months to initiate ART because of eligibility requirements and extensive preparation prior to initiation of therapy [23]. To ensure we captured these earlier CD4 cell count measurements, we widened the time window to six months before ART start to assess the percentage of adults with any CD4 cell count before ART start. If an adult had multiple CD4 cell count measurements within a time window, we selected the CD4 cell count measurement closest to ART start.

WHO recommends viral load testing six months after ART start; however in some parts of Southern Africa regional guidelines recommend the first viral load test at three months for pregnant and breastfeeding women, and at four months for adults aged ≥15 years [24]. We therefore assessed the percentage of adults with a viral load test three to nine months after ART initiation to ensure we captured all recommended testing periods and to allow up to a 3-month delay in testing. Among those with a viral load test, we calculated the percentage with an unsuppressed viral load (>1000 HIV-1 RNA copies/ml). To allow sufficient follow-up time to undertake viral load testing, we excluded adults who started ART less than nine months before database closure from the analysis of viral load testing. We also excluded adults who started ART in 2018 because of the small sample size; the analysis of viral load testing thus covered adults starting ART up to December 31, 2017 and viral load testing data up to nine months thereafter. If multiple viral load measurements occurred within the time window, we selected the first eligible viral load measurement.

### Statistical methods

We used descriptive statistics to summarize baseline characteristics at ART initiation, including percentage female, median age, median follow-up time and year of ART start stratified by country. We also used descriptive statistics to characterize trends in CD4 and viral load testing frequency and the proportion of adults with each test stratified by country. We used a mixed effect logistic regression model with random intercepts by country to assess the probability of having a CD4 cell count at ART start, a viral load test after ART start, and among those with a test, the probability of having advanced HIV disease or unsuppressed viral load over calendar year adjusted for age and sex. We created three age categories: <25, 25-49 and >49 years of age. We defined sex as either male or female and excluded 111 patients with unknown sex or date of birth. All statistical analyses were performed using Stata version 15.1 (Stata Corp, College Station, TX, USA).

### Ethical considerations

Local review boards and ethics committees approved the use of the data included in this study. The Cantonal Ethics Committee of the Canton of Bern, Switzerland, approved data merging and the collaborative analyses. Local review boards and the Cantonal Ethics Committee of the Canton of Bern waived the requirement to obtain informed consent.

## Results

Our main analysis included 502,456 adults from 14 programs encompassing more than 300 clinics across six countries in Southern Africa (Table 1). Median (IQR) age at ART initiation was 34.3 (28.6-41.3) years and 63.8% of patients were female. Median (IQR) follow-up time was 59.1 (28.8-95.3) months, ranging from 39.6 (20.6-60.7) months in Mozambique to 74.5 (39.3-107.8) months in South Africa. Most adults initiated ART in years 2009-2013 (43.2%), followed by years 2014-2018 (38.4%), and 2005-2008 (18.4%). The analysis of viral load testing after ART start included 458,528 adults (91.3% of adults included in the CD4 count analysis). Patient characteristics were similar but, as expected as a result of the broader exclusion criteria, the median follow-up time was longer (65.1 months) than in the CD4 count analysis (59.1 months) (Supplementary Table 1).

**Table 1.**
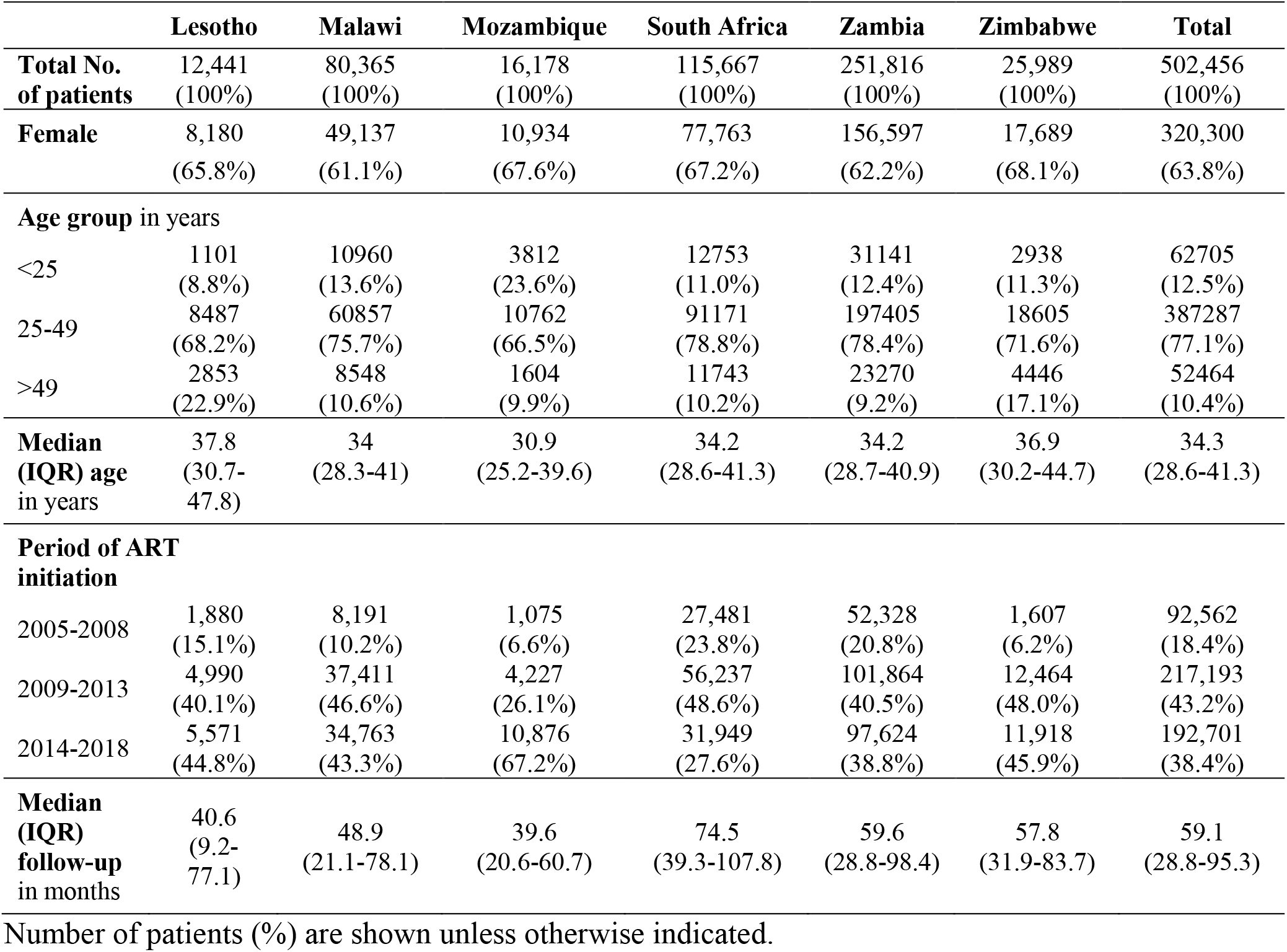
Characteristics of adult patients at antiretroviral therapy (ART) initiation by country 2005-2018.

### Frequency of CD4 cell count and viral load testing

There was an increase in the frequency of CD4 testing alongside an increasing number of adults in care in earlier years (Figure 1). However, in all countries the frequency of CD4 testing plateaued or declined, typically after 2010, despite an increasing number of adults in care and under follow-up in subsequent years; Malawi and South Africa had the strongest declines in testing. Figure 2 shows the frequency of viral load testing. In South Africa, viral load testing increased steeply from 2008 onwards but plateaued after 2012 (Figure 2). The other countries had little or no viral load testing prior to 2012. In Zambia, viral load testing increased sharply in 2015. Malawi experienced an earlier, but more gradual increase. In recent years, viral load testing also increased in Lesotho, Mozambique and Zimbabwe (Figure 2). The ratio of CD4 to viral load testing in the region decreased from a high of 7.2 in 2010 to a low of 2.1 in 2017 (Figure 3). The ratio decreased in each of the six countries (Supplementary Figure 1).

**Figure 1.**
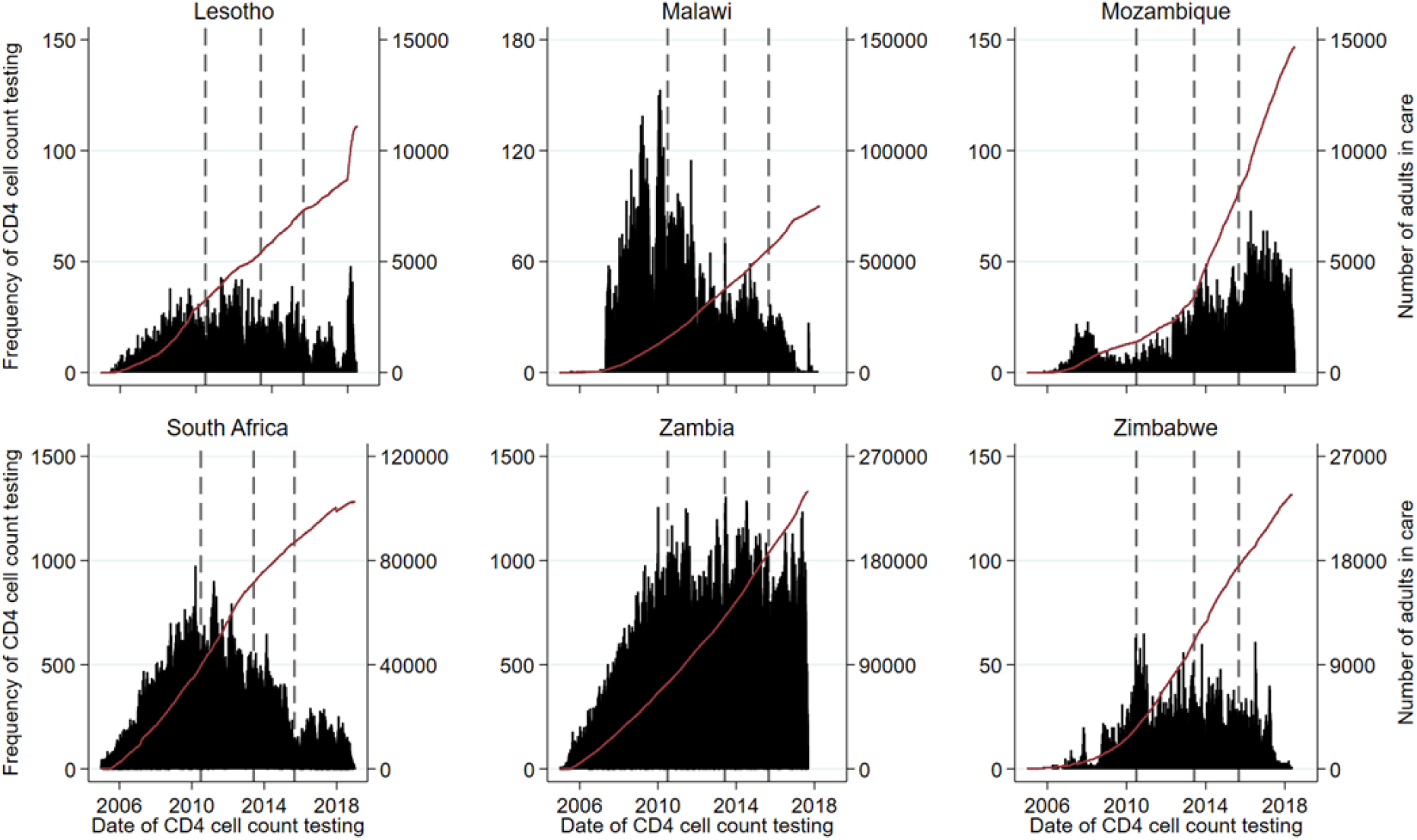
Frequency of CD4 cell count testing per day and cumulative number of adults in care by country. The vertical lines indicate the change in WHO guidelines. Note: scale of y-axis differs across countries.

**Figure 2.**
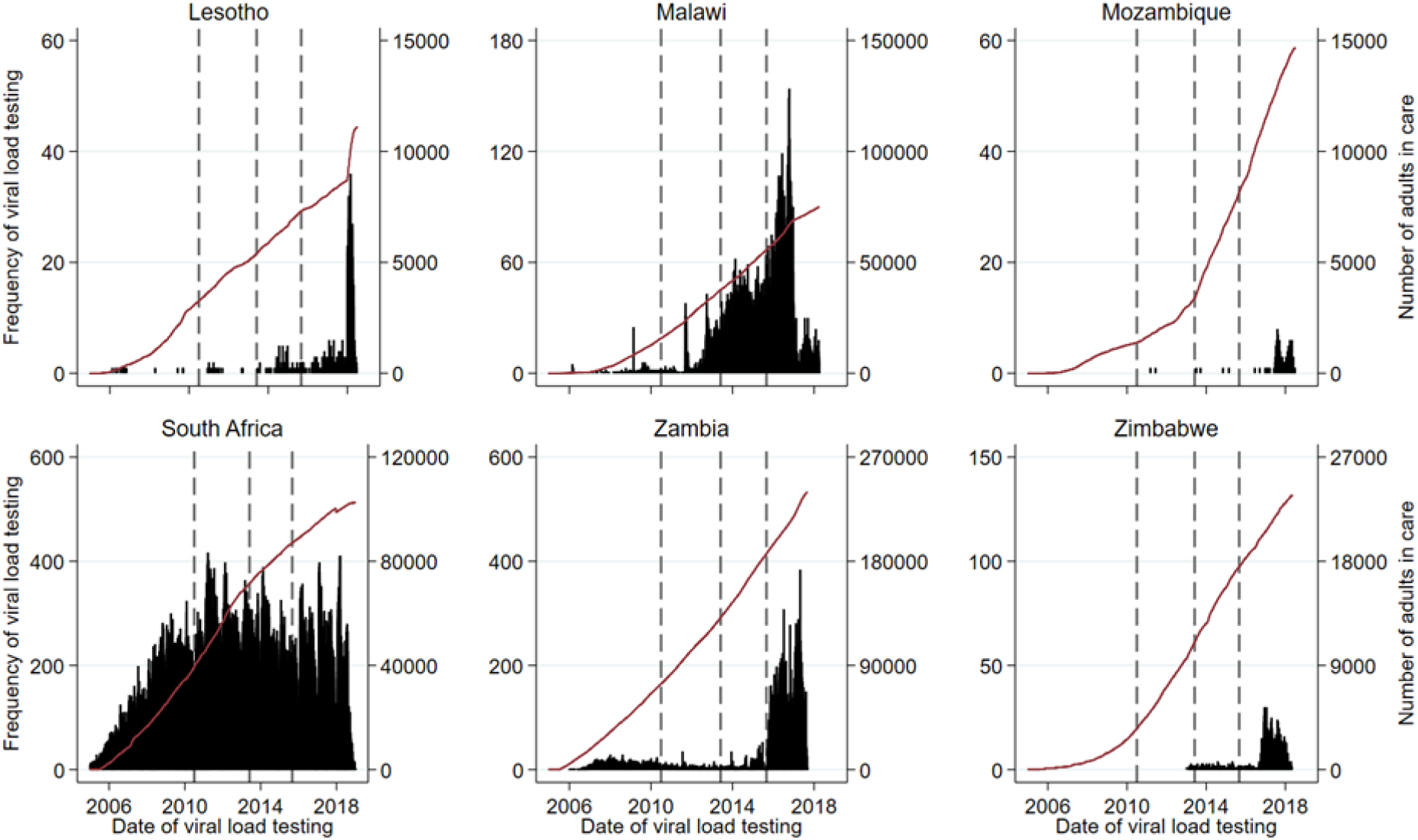
Frequency of viral load testing and number of adults in care by country. The vertical lines indicate the change in WHO guidelines. Note: scale of y-axis differs across countries.

**Figure 3.**
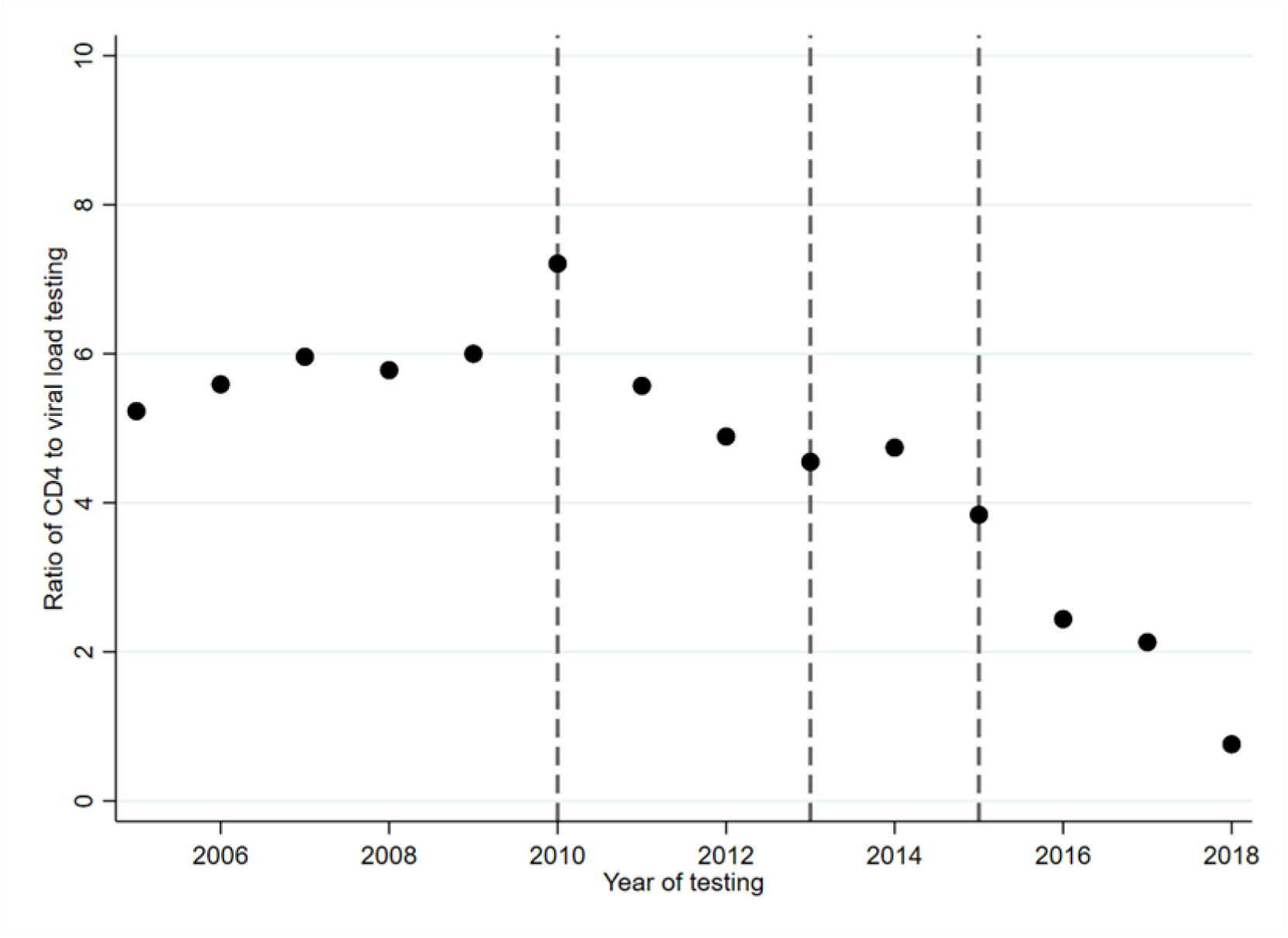
Trend of the ratio of CD4 cell count testing to viral load testing among adults. The vertical lines indicate the change in WHO guidelines.

### CD4 counts at ART initiation

Overall, 61.4% of adults had a CD4 cell count at ART start, ranging from 31.0% in Malawi to 81.4% in South Africa (Table 2). The percentage of adults with a CD4 cell count at ART initiation decreased over the years from a peak of 78.1% in 2008 to a low of 38.0% in 2017 (Figure 4), with an increase to 52.2% in 2018. Among those with a CD4 cell count at ART start, 53.6% had advanced HIV disease (Table 2). South Africa had the highest percentage of adults with advanced HIV disease (59.7%) and Mozambique the lowest (34.1%). The percentage of adults with advanced HIV disease steadily declined over time from a high of 83.3% in 2005 to a low of 23.5% in 2018 (Figure 4). Crude trends of the percentage of adults with a CD4 cell count test at ART start and, among those with CD4 testing, the percentage with advanced HIV disease by country are shown in Supplementary Figure 2. The probability of having a CD4 cell count at ART start was 14% lower (odds ratio [OR] 0.86; 95% CI 0.86-0.86) and the odds of having advanced HIV disease 20% lower each subsequent year (OR 0.80; 95% CI 0.80-0.81) (Table 3). Women had a lower probability of CD4 cell count testing at ART start (OR 0.86; 95% CI 0.85-0.87) and a lower probability of advanced HIV disease (OR 0.63; 95% CI 0.62-0.64) than men (Table 3). Adults aged 25 years or older had a higher probability of CD4 cell count testing at ART start than adults under 25 years of age. Among those with testing, adults under 25 years of age had a lower probability of advanced HIV disease than adults 25 years of age or older.

**Table 2.** CD4 cell count testing at antiretroviral therapy start (ART) and viral load testing after ART start.

|  | Lesotho | Malawi | Mozambique | South Africa | Zambia | Zimbabwe | Total |
| --- | --- | --- | --- | --- | --- | --- | --- |
| **CD4 cell count at ART start** |  |  |  |  |  |  |  |
| Total No. of patients | 12,441<br>(100%) | 80,365<br>(100%) | 16,178<br>(100%) | 115,667<br>(100%) | 251,816<br>(100%) | 25,989<br>(100%) | 502,456<br>(100%) |
| Patients with CD4 cell count testing at ART start | 9,230<br>(74.2%) | 24,925<br>(31.0%) | 10,472<br>(64.7%) | 94,167<br>(81.4%) | 155,459<br>(61.7%) | 14,199<br>(54.6%) | 308,452<br>(61.4%) |
| Patients with advanced HIV disease among those with CD4 cell count testing at ART start | 3,777<br>(40.9%) | 1,301<br>(52.2%) | 3,570<br>(34.1%) | 56,228<br>(59.7%) | 81,458<br>(52.4%) | 7,249<br>(51.1%) | 165,278<br>(53.6%) |
| **Viral load testing after ART start** |  |  |  |  |  |  |  |
| Total No. of patients | 9,386<br>(100%) | 70,887<br>(100%) | 14,698<br>(100%) | 110,520<br>(100%) | 228,699<br>(100%) | 24,338<br>(100%) | 458,528<br>(100%) |
| Patients with viral load testing 3-9 months after ART start | 41<br>(0.4%) | 7,574<br>(10.7%) | 17<br>(0.1%) | 67,409<br>(61.0%) | 4,222<br>(1.9%) | 195<br>(0.8%) | 79,458<br>(17.3%) |
| Patients with unsuppressed viral load among those with viral load testing 3-9 months after ART start | 12<br>(29.3%) | 646<br>(8.5%) | 12<br>(70.6%) | 5,700<br>(8.5%) | 442<br>(10.5%) | 31<br>(15.9%) | 6,843<br>(8.6%) |

Number of patients (%) are shown.
Because of the shorter study period, the number of patients included in the analyses of viral load testing after antiretroviral therapy (ART) start is smaller (n=458,528) than the number included in the analysis of CD4 cell count testing at ART start (n=502,456).
Advanced HIV disease defined as CD4 cell count <200 cells/mm<sup>3</sup>; unsuppressed viral load defined as HIV-1 RNA >1000 copies/ml.

**Table 3.** Probability of CD4 cell count testing at antiretroviral therapy (ART) start and viral load testing after ART start.

|  | CD4 cell count at<br>ART start<br>N=502,456 | Advanced HIV<br>disease<br>N=308,452 | Viral load after<br>ART start<br>N=458,528 | Unsuppressed<br>viral load<br>N=79,458 |
| --- | --- | --- | --- | --- |
|  | OR (95% CI) | OR (95% CI) | OR (95% CI) | OR (95% CI) |
| **Per calendar**<br>**year** | 0.86<br>(0.86-0.86) | 0.80<br>(0.80-0.81) | 1.06<br>(1.05-1.06) | 1.00<br>(0.99-1.01) |
| **Age group in**<br>**years** |  |  |  |  |
| <25 | 1 | 1 | 1 | 1 |
| 25-49 | 1.18<br>(1.16-1.21) | 1.50<br>(1.47-1.54) | 1.45<br>(1.40-1.50) | 0.70<br>(0.65-0.76) |
| >49 | 1.20<br>(1.17-1.24) | 1.48<br>(1.43-1.53) | 1.34<br>(1.28-1.41) | 0.52<br>(0.46-0.59) |
| **Sex** |  |  |  |  |
| Male | 1 | 1 | 1 | 1 |
| Female | 0.86<br>(0.85-0.87) | 0.63<br>(0.62-0.64) | 1.06<br>(1.04-1.09) | 0.79<br>(0.75-0.83) |

Results from the mixed-effects logistic regression model showing the probability of having a CD4 cell count at antiretroviral therapy (ART) start, a viral load test after ART start and, among those with testing, the probability of having advanced HIV disease and unsuppressed viral load by calendar year, age and sex.
CD4 cell count analyses based on 502,456 adults of whom 308,452 (61.4%) had CD4 cell count testing at ART start; viral load analyses based on 458,528 adults of whom 79,458 (17.3%) had viral load test testing after ART start.
Advanced HIV disease defined as CD4 cell count <200 cells/mm<sup>3</sup>; unsuppressed viral load defined as HIV-1 RNA >1000 copies/ml.

**Figure 4.**
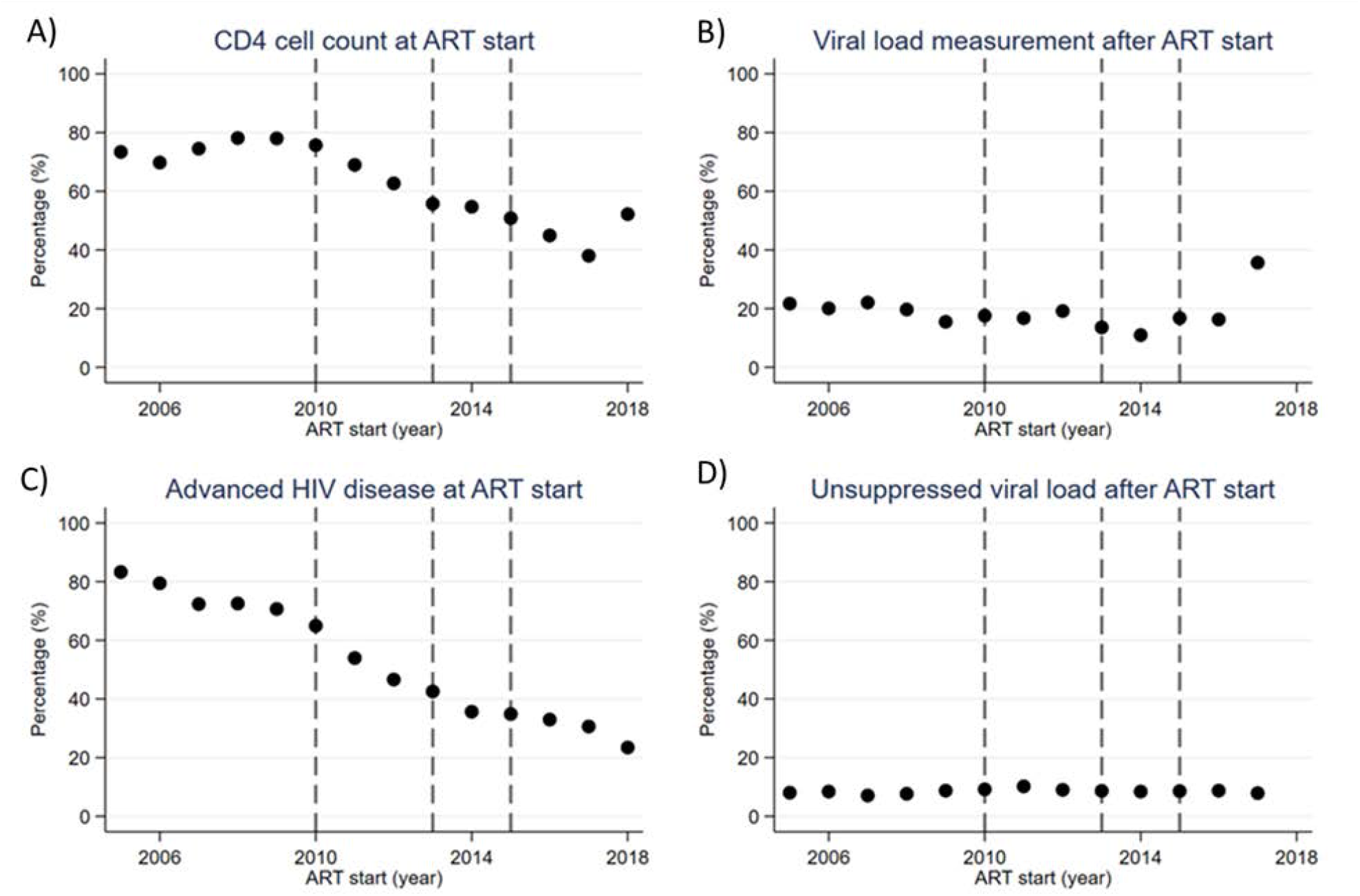
Trends of CD4 cell count testing at antiretroviral therapy (ART) start and viral load testing after ART start. (A) The percentage of patients with a CD4 cell count at antiretroviral therapy (ART) start and, among those, (C) the percentage with advance HIV disease; and (B) the percentage with a viral load test after ART start and, among those, (D) the percentage with unsuppressed viral load by year of ART start. Advanced HIV disease defined as CD4 cell count <200 cells/mm3; unsuppressed viral load defined as HIV-1 RNA >1000 copies/ml. The vertical lines indicate the change in WHO guidelines.

Expanding the CD4 cell count testing time window from three to six months before ART initiation increased the percentage of adults with a CD4 cell count at ART start, with the difference deceasing over time (Supplementary Figure 3).

### Viral load testing on ART

Overall, 17.3% of adults had a viral load test three to nine months after ART start, among whom 8.6% were unsuppressed (Table 2). South Africa had the highest percentage of adults with viral load testing after ART start (61.0%), followed by Malawi (10.7%); viral load testing was below 2% in all other countries. Among those with viral load testing, the percentage with unsuppressed viral load was lowest in South Africa and Malawi (8.5% and 8.5%, respectively) and highest in Mozambique (70.6%). Crude trends of the percentage of adults with a viral load test after ART start and the percentage with unsuppressed viral load by country are shown in Supplementary Figure 2.

In years 2005 to 2016, 10% to 22% of adults had a viral load test after ART start, which increased to nearly 36% in 2017 (Figure 4). The mixed effect logistic regression model showed that the increase over time in the probability of having a viral load test was modest (OR 1.06; 95% CI 1.05-1.06). There was no evidence of a decrease in unsuppressed viral load over time (OR 1.00; 95% CI 0.99-1.01) (Table 3). Overall, the percentage with unsuppressed viral load consistently remained in the range of 7% to 10% across all years (Figure 4). The probability of having a viral load measurement at ART start was higher for adults aged ≥25 years, and, among those with a test, the probability of having an unsuppressed viral load was lower than for adults aged <25 years (Table 3). Women had a slightly higher probability of having a viral load test after ART start (OR 1.06; 95% CI 1.04-1.09) and a lower probability of having an unsuppressed viral load (OR 0.79; 95% CI 0.75-0.83) than men.

## Discussion

This large study of CD4 and viral load testing in ART programs from six southern African countries showed that the percentage of adults with a CD4 cell count at the start of ART declined from about 80% in 2005 to below 40% in 2017. Among those with a CD4 count, the percentage starting with advanced HIV disease (<200 cells/mm^3^) also declined, but almost a quarter of patients still started ART with advanced disease in 2018. Viral load testing increased in the earlier years in South Africa and then plateaued. In the other countries testing increased in more recent years, but remained modest considering the large number of patients in care. In 2017, overall only about a third of patients had a viral load measurement after the start ART.

The trends in CD4 testing observed in this study likely reflect changes in ART guidelines. In earlier years, when ART eligibility was governed by relatively low CD4 thresholds (<200 cells/mm^3^ up to 2010 [25], ≤350 up to 2013 [26,27]), repeated CD4 counts were needed to identify when patients became eligible for ART. As eligibility expanded to higher CD4 thresholds and, in September 2015, “Treat all” guidance was introduced recommending that all individuals be treated as soon as possible after diagnosis of HIV infection, less CD4 testing was required [28]. Of note, the peak in the ratio of CD4 to viral load testing in 2010 coincides with the release of the guideline recommending lifelong ART for pregnant women under Option B [12,29]. In the earlier years, WHO and national guidelines recommended routine CD4 cell count monitoring for patients on ART [27,30]. However, in 2012 the Southern African HIV Clinicians Society released new guidelines recommending viral load monitoring rather than routine CD4 cell count monitoring for patients on ART with a CD4 cell count above 200 cells/mm^3^ [31]. The following year, WHO and the South African Department of Health released updated guidelines that no longer recommended routine CD4 cell count monitoring for patients on ART where viral load monitoring was assured [12,32].

The decline in CD4 cell count testing at the start of ART in the six countries included in this analysis is of concern. Although CD4 cell count thresholds for starting ART and CD4 cell count monitoring on ART are no longer recommended in the “Treat all” era, WHO still recommends baseline CD4 cell count testing for all patients entering care and before ART initiation to identify patients with advanced HIV disease [5,7]. CD4 cell count testing remains important because clinical staging has limited diagnostic accuracy for identifying patients with low CD4 cell count [33]. WHO recommends a package of interventions among patients with advanced HIV disease, including screening, treatment and/or prophylaxis for major opportunistic infections, rapid ART initiation and intensified adherence support [8]. Scaling up access to this package of interventions to reduce mortality and morbidity is also being supported by major donors such as UNITAID [34]. The decrease in the proportion of patients with a CD4 cell count at the start of ART indicates that many patients with advanced HIV disease may be missed. The percentage with a CD4 count increased in 2018 to 52.2%. Future analyses of the IeDEA cohorts will examine whether or not this reversal of the trend is sustained.

PEPFAR, which provided substantial support to all countries included in this study, recently shifted their policy away from routine baseline CD4 cell count testing to prioritize access to viral load monitoring [16,17]. PEPFAR continues to support limited CD4 testing in countries where more than 10% of patients start ART with advanced HIV disease [16,17]. We found that almost a quarter of patients with a CD4 count started ART with advanced disease in 2018, but many started without a CD4 cell count. A recent multiregional analysis of IeDEA data used multiple imputation to adjust for missing CD4 cell counts and found that although median CD4 cell counts at ART start increased substantially from 2002 to 2015, the proportion with advanced disease (CD4 cell count <200 cells/mm^3^) was around 40% in low-income and middle-income countries in 2015 [35]. It is likely that the proportion of patients with advanced HIV disease at the time of initiating ART is above the PEPFAR threshold of 10%, indicating the need for continued baseline CD4 cell count testing.

Viral load testing is the preferred method for monitoring patients on ART, as treatment failure is identified earlier and more accurately than CD4 cell count monitoring [5,36]. Despite the recommendation to switch from CD4 to viral load monitoring, a substantial increase in viral load testing parallel to the decrease in CD4 count testing was evident only in South Africa and Malawi. In the other four countries included in this study, Lesotho, Mozambique, Zambia and Zimbabwe, the scale-up of viral load testing started only very recently. The higher rate of unsuppressed viral load in these latter countries further indicates they are still transitioning from targeted to routine viral load testing.

Two major strengths of this study are the timeliness of data and the large number of patients: we included data reported up to 2018 on more than 500,000 people living with HIV in six countries in Southern Africa. Our study also has several limitations. The ART programs and clinics participating in IeDEA Southern Africa may not represent all treatment facilities located in the region; therefore, the findings in this study may not be applicable to the country or region as a whole. Our study did not account for differences in level of care or access to laboratory testing that may vary across and within countries and be a result of different facility type and location. Our study focused on patients who started ART and did not consider patients lost to follow-up before or after ART, patients who silently transferred to another ART program or unreported mortality. Lastly, we were unable to ascertain trends in CD4 cell count testing among patients with suspected treatment failure, which is recommended by WHO and many national guidelines. Although this study analysed recently collected data, the findings will not reflect the most recent policy changes.

## Conclusions

This large multi-cohort study of people living with HIV in Southern Africa showed that CD4 cell count testing declined over time, including testing at the start of ART, despite the fact that many patients still initiate ART with advanced HIV disease. Although guidelines and national policies support the scale-up of viral load testing, the coverage of viral load testing was low in most countries. Without CD4 cell count testing and expanded viral load testing many patients with advanced HIV disease and treatment failure may go undetected, threatening the effectiveness of ART in southern Africa.

## Data Availability

Complete data for this study cannot be posted in a supplemental file or a public repository because of legal and ethical restrictions. Disclosure of a person’s HIV status can be highly stigmatizing, and since reidentification of deidentified datasets may be possible when they are combined with publicly available datasets (see work of Dr. Latanya Sweeney), IeDEA promotes the signing of a Data Use Agreement before HIV clinical data can be released. To request data, readers may contact IeDEA for consideration and instructions by filling out the online form available at www.iedea.org/home/who-we-are and completing the application at www.iedea.org/wp-content/uploads/2017/05/IeDEA_Multiregional_Concept_Application_Form_August_2016.docx

## Authors’ Contributions

EZ, ME, NF, FC, MAD, JVD, JE and MPF conceptualized the study, while EZ, CHDO, NM, MAD, JE, JVD, SB, SP, FT, NS, JM, GF, HP, RW, MPF and ME contributed to data collection. EZ, FC and ME analysed the data and interpreted the results. EZ wrote the first draft of the manuscript, which was revised by all authors. All authors have reviewed and approved the final version of the manuscript.

## Site investigators and cohorts

Gary Maartens, Aid for AIDS, South Africa; Carolyn Bolton, Centre for Infectious Disease Research in Zambia (CIDRZ), Zambia; Robin Wood, Gugulethu ART Programme, South Africa; Nosisa Sipambo, Harriet Shezi Clinic, South Africa; Frank Tanser, Africa Centre for Health & Population Studies (Hlabisa), South Africa; Andrew Boulle, Khayelitsha ART Programme, South Africa; Geoffrey Fatti, Kheth’Impilo, South Africa; Sam Phiri, Lighthouse Clinic, Malawi; Cleophas Chimbetete, Newlands Clinic, Zimbabwe; Karl Technau, Rahima Moosa Mother and Child Hospital, South Africa; Brian Eley, Red Cross Children’s Hospital, South Africa; Josephine Muhairwe, SolidarMed Lesotho; Juan Burgos-Soto, SolidarMed Mozambique; Cordelia Kunzekwenyika, SolidarMed Zimbabwe, Matthew P Fox, Themba Lethu Clinic, South Africa; Hans Prozesky, Tygerberg Academic Hospital, South Africa.

## Data centers

Nina Anderegg, Marie Ballif, Lina Bartels, Julia Bohlius, Benedikt Christ, Felix Cuneo, Cam Ha Dao Ostinelli, Masa Davidovic, Tafadzwa Dhokotera, Matthias Egger, Lukas Fenner, Andreas Haas, Anthony Hauser, Stefanie Hossmann, Serra Lem, Catrina Mugglin, Martina Reichmuth, Julien Riou, Veronika Skrivankova, Lilian Smith, Katayoun Taghavi, Per von Groote, Gilles Wandeler, Elizabeth Zaniewski, Kathrin Zürcher, Institute of Social and Preventive Medicine, University of Bern, Switzerland; Kim Anderson, Andrew Boulle, Morna Cornell, Mary-Ann Davies, Victoria Iyun, Leigh Johnson, Reshma Kassanjee, Kathleen Kehoe, Mmamapudi Kubjane, Nicola Maxwell, Ernest Mokotoane, Patience Nyakato, Gem Patten, Priscilla Tsondai, Renee de Waal, School of Public Health and Family Medicine, University of Cape Town, South Africa.

## Supporting Information

**Appendix 1:** Supplementary material.

**Supplementary Table 1**: **Characteristics of patients at antiretroviral therapy (ART) initiation for the sub-analysis of viral load testing after ART start by country**.

Number of patients (%) are shown unless otherwise indicated. SuppInfo_Table_1.docx

**Supplementary Figure 1**: **Trends of the ratio of CD4 cell count testing to viral load testing by country**.

The vertical lines indicate the change in WHO guidelines. N/A: no CD4 cell count or viral load testing data available for patients who started ART in that year. Advanced HIV disease defined as CD4 cell count <200 cells/mm^3^; unsuppressed viral load defined as HIV-1 RNA >1000 copies/ml. SuppInfo_Figure_1.docx

**Supplementary Figure 2**: **Trends of CD4 cell count testing at antiretroviral therapy (ART) start and viral load testing after ART start by country**.

The percentage of patients with a CD4 cell count at initiation of antiretroviral therapy (ART) and, among those, the percentage with advanced HIV disease; and the percentage with a viral load test after ART start and, among those, the percentage with unsuppressed viral load. The vertical lines indicate the change in WHO guidelines. N/A: no CD4 cell count or viral load testing data available for patients who started ART in that year. Advanced HIV disease defined as CD4 <200 cells/mm^3^; unsuppressed viral load defined as measurement HIV-1 RNA >1000 copies/ml. SuppInfo_Figure_2.docx

**Supplementary Figure 3**: **Trends of CD4 cell count testing three months vs six months before antiretroviral therapy start**.

The percentage of patients with a CD4 cell count up to three months before antiretroviral therapy (ART) start and up to six months before ART start and, among those with testing, the percentage with advanced HIV disease. The vertical lines indicate the change in WHO guidelines. Advanced HIV disease defined as CD4 <200 cells/mm^3^.

SupplInfo_Figure_3.docx

